# Sociodemographic and Mental Health Predictors of Mental Health Service Use Across Provider Types

**DOI:** 10.1101/2025.06.03.25328937

**Authors:** Nelson Pang, Jessie Yeung

## Abstract

**Objective:** To examine trends in mental health service use across four provider types (family doctors, psychiatrists, psychologists, and social workers) and identify sociodemographic predictors of provider-specific access in Canada

**Methods:** This study used data from seven cycles (2007–2020) of the Canadian Community Health Survey (CCHS), a nationally representative cross-sectional survey. Trends over time were examined using weighted proportions and counts of service users. Multivariable logistic regression models were conducted on the 2019–2020 cycle to assess associations between sociodemographic factors and provider-specific service use. Analyses were weighted and adjusted for sex, age, income, education, visible minority status, immigrant status, Aboriginal identity, and self-perceived health.

**Results:** Family doctors were consistently the most accessed providers for mental health concerns, followed by psychologists and social workers, with psychiatrists being least accessed. Psychologist and social worker use increased between 2017 and 2019. In regression models, women had significantly higher odds of using family doctors and social workers, while older adults had greater odds of accessing family doctors and lower odds of accessing psychologists and social workers. Higher education was associated with increased psychologist use, whereas lower income and education were linked to family doctor and social work use. Non-Indigenous individuals had higher odds of seeing a psychologist, and those not identifying as a visible minority had higher odds of seeing a family doctor. Poorer perceived mental health was a strong predictor of accessing all provider types.

**Conclusions:** This study reveals a stratified mental health care system in Canada, where sociodemographic factors shape who accesses which providers. While primary care dominates, growth in psychologist and social worker use suggests shifting patterns of engagement. Findings underscore the need for policies that address financial and structural barriers, promote equitable access, and expand coverage for community-based mental health providers.

## Background

Mental health is a significant public health concern in Canada, with approximately 1 in 5 Canadians experiencing a mental illness every year.^1^ Despite this access to mental health care is limited with studies estimating that only around 40–60% of people with a mental health problem seeking professional assistance.^2,3^ In 2018, an estimated 5.3 million Canadians reported needing help for their mental health in the previous year highlighting the widespread demand for mental health support across the Canadian population.^4^ Alarmingly, nearly half of these individuals had their needs only partially met (1.2 million) or completely unmet (1.1 million), suggesting substantial gaps in service delivery for mental health. ^4^

In Canada, mental health care is provided by a diverse range of professionals, including psychiatrists, psychologists, social workers, nurses, and physicians.^3^ These different providers differ not only in their training and scope of practice but also in funding and accessibility. For example, family physicians and psychiatrists are typically publicly funded and can be billed directly to provincial and territorial health insurance plans whereas services provided by psychologists and social workers may require out-of-pocket payment or private insurance coverage. These differences in cost of services may result in financial barriers for individuals seeking care. Furthermore, these differences have implications on who receives care, what kind of care is delivered, and how barriers to access may differ across populations. In recent years, the mental health service landscape in Canada has become increasingly complex with expanding roles in the mental health care for non-physician providers such as psychologists and social workers.^5^ This change reflects both efforts to meet growing demand for mental health care, shifts toward evidence-based care, and interprofessional models of service delivery.^6,7^

Previous research has shown that mental health care utilization is shaped by an interplay of need, enabling factors and predisposing factors, such as symptom severity, income, gender, education, cultural background, wait time, and costs.^8–12^ Despite Canada having universal health insurance, disparities and inequities are still associated with the underutilization of mental health care services.^8,11,13,14^ For example, previous research in Canada has found that individuals with higher education are more likely to utilize mental health services than those in less educated groups.^11,13^ Likewise, previous research has found income-based inequity in access to mental health services.^8,14^ Understanding which populations access specific types of providers is critical for informing mental health care and equity-focused policies. For example, investments in community-based providers such as psychologists and social workers may reduce reliance on primary care for mental health, improve continuity of care, and expand culturally appropriate service options.^15–17^

A limitation in this research however is that most studies aggregate all types of mental health providers into one variable limiting for the examination of differences in who accesses what services and under what circumstances. Additionally, because of this aggregation there are limited studies that examine trends overtime on how Canadians engage with different mental health care providers. This study focuses on examining how sociodemographic and mental health characteristics are associated with mental health service use across four provider types (family doctors, psychiatrists, psychologists, and social workers) and trends in their use over time in Canada. This research will addresses key knowledge gaps by disaggregating provider-specific service use and examine how these patterns vary across populations and years.

## Methods

### Data Source

This study draws from seven cycles of the Canadian Community Health Survey (CCHS) collected between 2007 and 2020. The CCHS is a nationally representative cross-sectional survey administered by Statistics Canada. The CCHS is a self-reported survey on health, healthcare utilization, and health determinants of the Canadian population aged 12 and above.

Sample sizes varied across cycles, and all analyses applied survey weights provided by Statistics Canada to ensure representativeness of the Canadian population.

### Ethics, Data Access, and Participant Confidentiality

This study used retrospective survey data from the CCHS Public Use Microdata Files (PUMFs), accessed for research purposes in January 2025. These data are collected and publicly released by Statistics Canada, which obtains informed consent from participants at the time of original data collection. The PUMFs are fully anonymized prior to release and do not contain any identifying information. As a result, the authors did not have access to any information that could identify individual participants during or after data collection, and additional institutional ethics approval was not required for this secondary analysis.

### Outcome variables

The data was subsetted to only include respondents who have consulted a mental health professional in the past 12-months (n=13,580/weighted n= 4,679,132.44). The primary outcome were binary indicators of if respondents reported having seen or talked to a specific health professional based on the question *“In the past 12 months have you seen or talked to a health professional about your emotional or mental health?”*. The analysis focused on the four most common providers for mental health care (family doctor, psychiatrist, psychologist, and social work). Each provider was coded as a separate variable as respondents could see more than one provider.

### Predictor variables

A variety of sociodemographic and mental health characteristics were included as predictors in the model. Sociodemographic characteristics included sex, age, household income, visible minority status, education, immigration status, and Aboriginal identity. Mental health characteristics included perceived health and perceived mental health.

### Statistical Analysis

An exploratory descriptive analysis was conducted to examine trends in mental health service use across provider types over time (2007-2020). To identify factors associated with provider-specific mental health service use four separate logistic regression models were conducted using the 2019-2020 cycle of the data for each provider type (family doctor, psychiatrist, psychologist, and social worker). Each model assessed the relationship between the outcome variables and the likelihood of using that provider adjusting for all other covariates.

Two-tailed tests with a threshold of α = 0.05 were used for all statistical tests. All analyses were conducted using R statistical software and survey weights were applied to account for the complex sampling design of the CCHS and to be representative of the Canadian population.

## Results

Between 2007 and 2020, family doctors consistently accounted for the highest proportion of mental health service use in Canada with over half of service users responding to having accessed a family doctor for mental health care (See Figure 1). The proportion of people who accessed care through psychologists and social workers showed a gradual increase over time, while the proportion of visits to psychiatrists remained relatively unchanged over time. In 2017, there was a significant increase in the proportion of those who accessed a psychologist potentially reflecting data issues in the 2017 cycle of the data.

**Figure 1.**
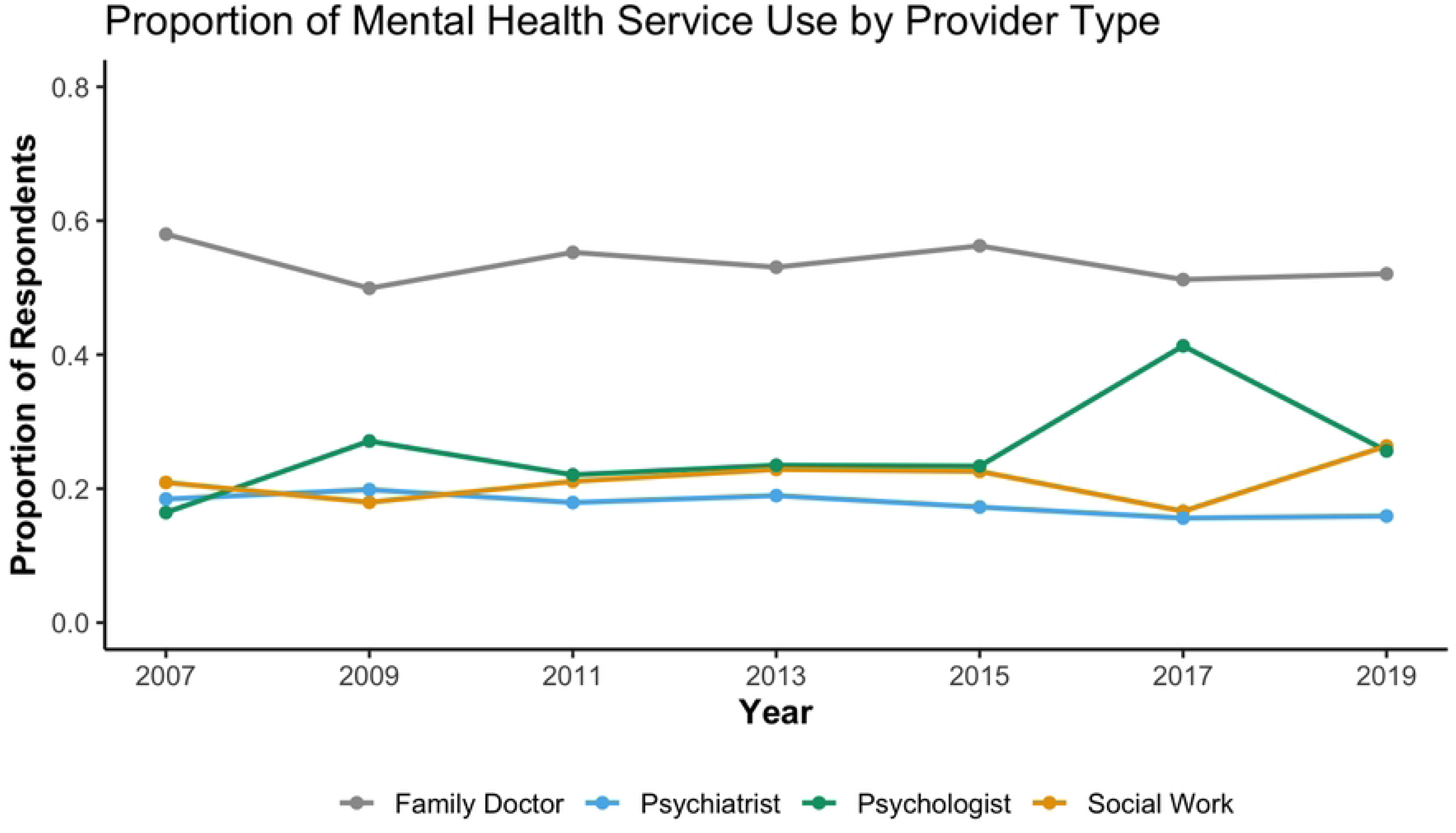
Trends in Proportion of Mental Health Service Use by Provider Type.

When examining weighted estimates of the number of Canadians accessing care, similar trends were observed (See Figure 2). The total number of individuals seeing family doctors for mental health reasons far exceeded those accessing psychologists, psychiatrists, or social workers. However, all provider types showed increases in absolute numbers over time reflecting increased mental service use overtime. Notably, a temporary drop in service use was observed in 2017 across all provider types, potentially reflecting data collection limitations in that survey cycle.

**Figure 2.**
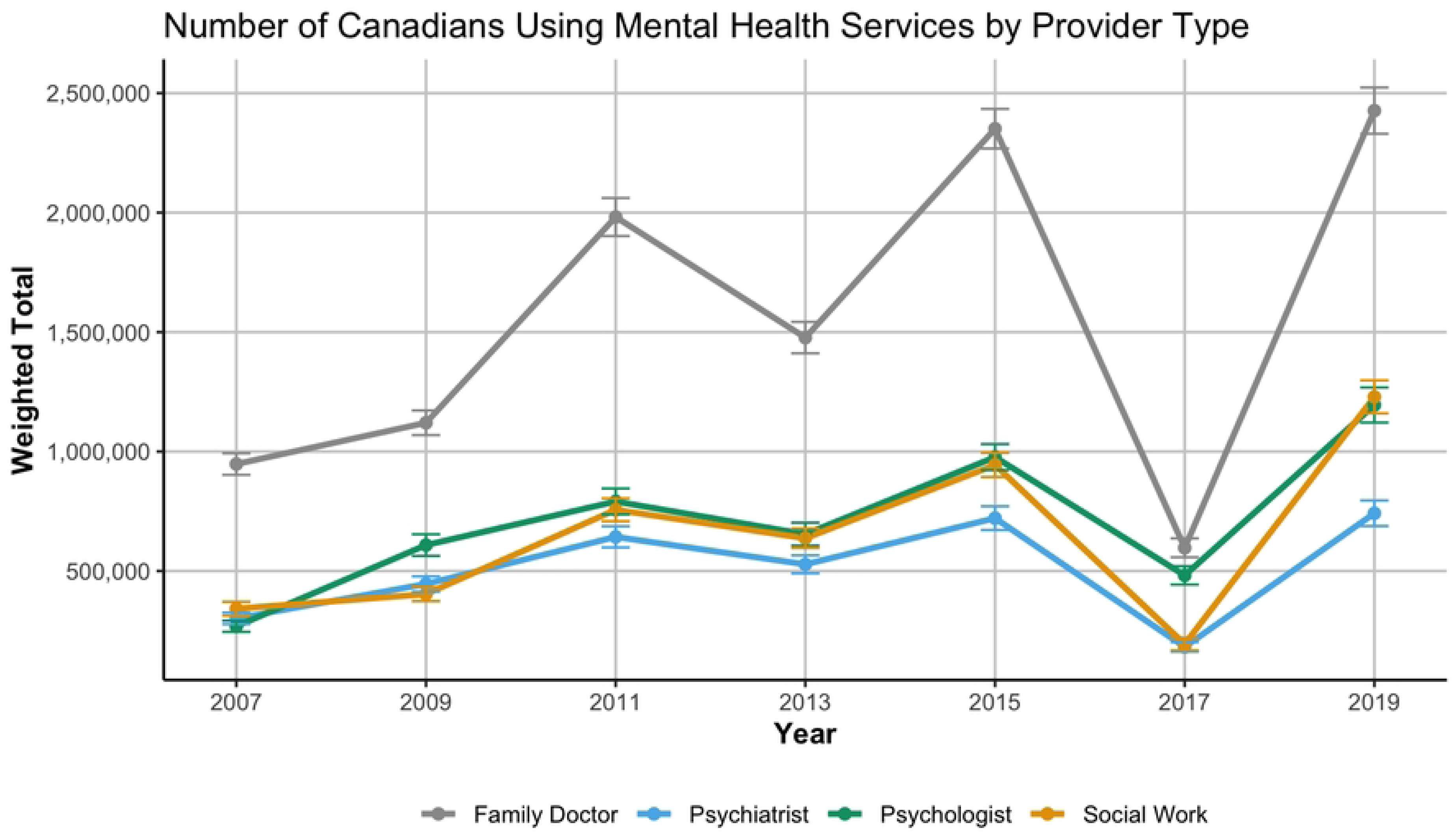
Weighted Number of Canadians Using Mental Health Services by Provider Type.

Associations between sociodemographic and mental health characteristics and provider-specific mental health service use are presented in Table 1. Women had significantly higher odds than men of using social work (OR = 1.19, 95% CI: 1.02–1.40, p = 0.028) and family doctor services (OR = 1.21, 95% CI: 1.05–1.39, p = 0.007), but lower odds of seeing a psychiatrist (OR = 0.66, 95% CI: 0.55–0.79, p < .001).

**Table 1.**
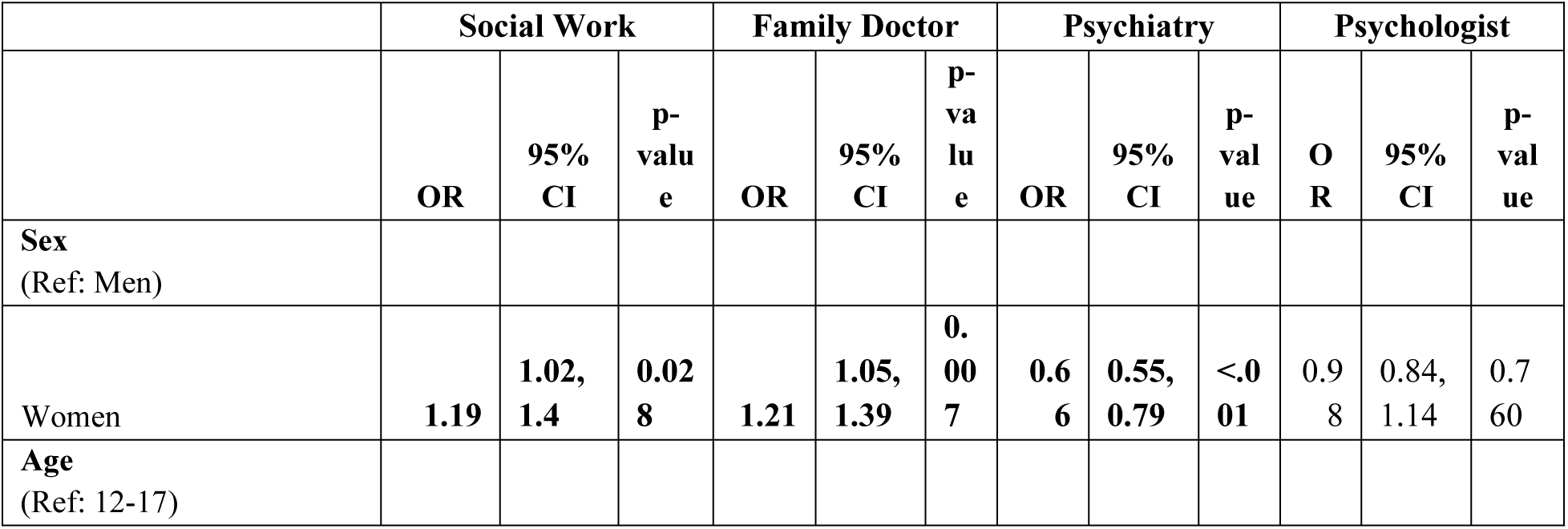
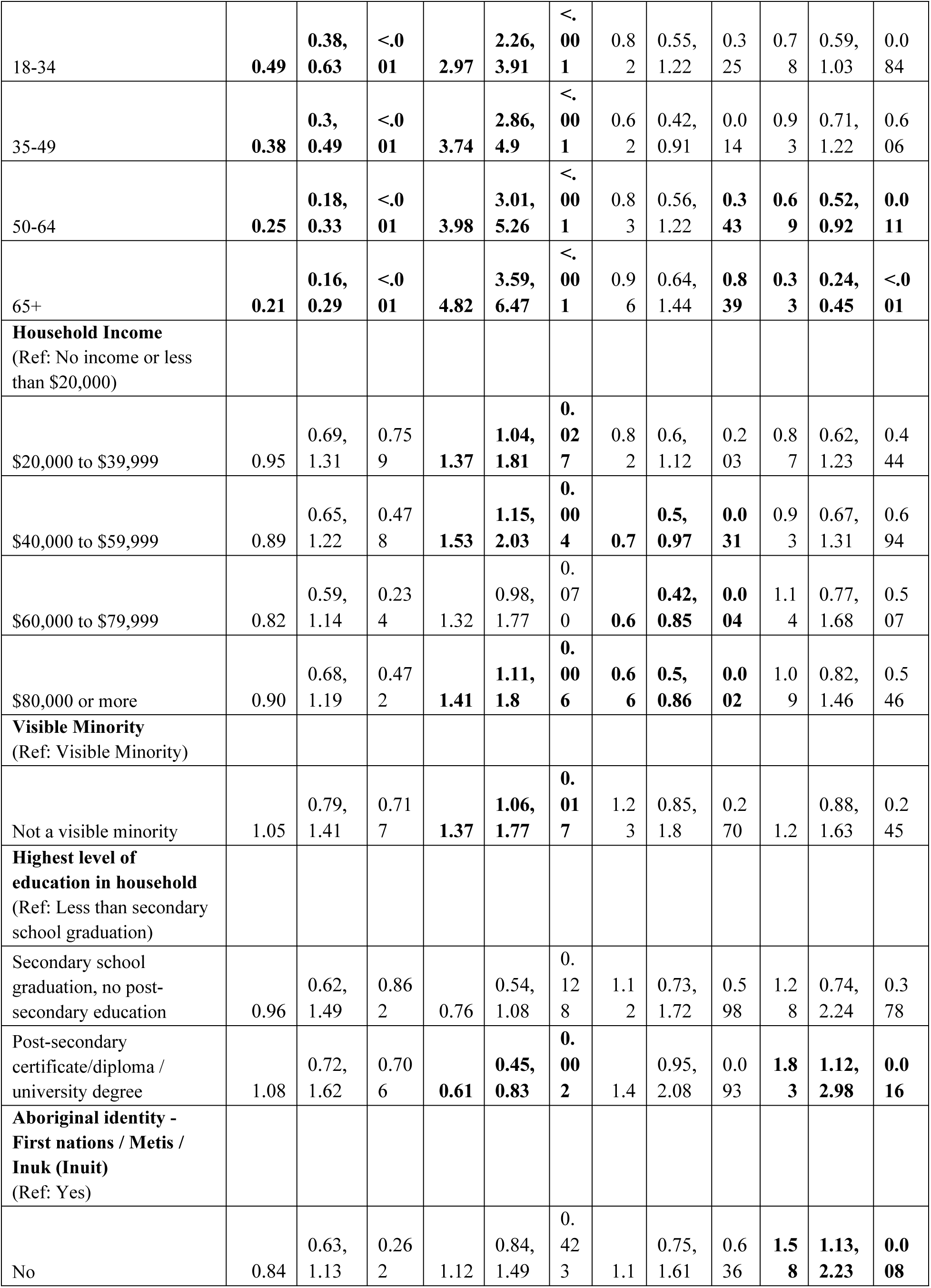
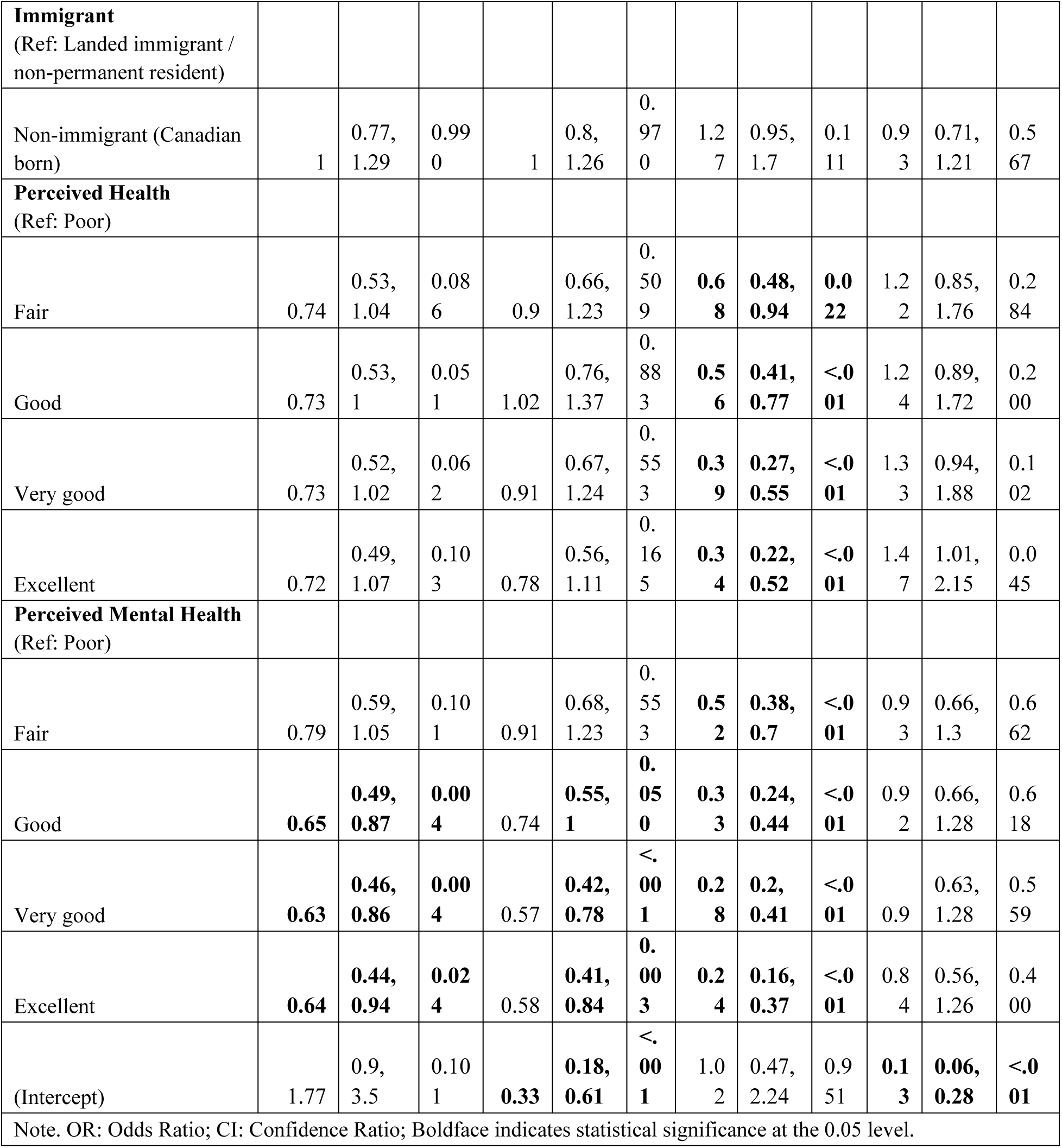
Associations between Sociodemographic Variables and Mental Health Service Provider Use.

Older age was associated with increased use of family doctors. Specifically, those age 65+ had the highest odds (OR = 4.82, 95% CI: 3.59–6.47, p < .001), but lowest odds of using social workers (OR = 0.21, 95% CI: 0.16–0.29, p < .001) and psychologists (OR = 0.33, 95% CI: 0.24–0.45, p < .001). For household income, individuals in all income groups over $40,000 had significantly lower odds of accessing psychiatrist. Notably, those in the highest income group ($80,000+) had lower odds of seeing a psychiatrist (OR = 0.66, 95% CI: 0.50–0.86, p = 0.002) compared to the lowest income group category. In contrast, individuals in higher income groups has higher odds of accessing a family doctor for mental health. Conspicuously, there were no significant associations found between income and psychologist use. Additionally, educational attainment was significantly associated with psychologist use. Compared to households with no high school attainment, respondents in households with a post-secondary degree had higher odds of accessing psychologists (OR = 1.83, 95% CI: 1.12–2.98, p = 0.016), but lower odds of accessing family doctors (OR = 0.61, 95% CI: 0.45–0.83, p = 0.002).

Few significant associations emerged for immigrant status or Indigenous identity, with the exception that non-Indigenous respondents had significantly higher odds of psychologist use (OR = 1.58, 95% CI: 1.13–2.23, p = 0.008). Similarly, not being a visible minority was associated with higher odds of seeing a family doctor (OR 1.37, 95% CI: 1.06-1.77, p=0.017) but no other provider use was significant.

Perceived mental health was a consistent and strong predictor of provider use. Individuals reporting “excellent” perceived mental health had significantly lower odds of seeing a psychiatrist (OR = 0.24, 95% CI: 0.16–0.37, p < .001), family doctor (OR = 0.58, 95% CI: 0.41–0.84, p = 0.003), or social worker (OR = 0.64, 95% CI: 0.44–0.94, p = 0.024), compared to those reporting poor mental health.

## Discussion

This study examined national trends in mental health service use across four provider types (family doctors, psychiatrists, psychologists, and social workers) and identified sociodemographic predictors to accessing each provider using a recent nationally representative survey. Results show that family doctors were the most common provider for mental health, followed by psychologists and social workers, while psychiatrists are accessed less frequently. These trends were generally stable over time, with increases in psychologist and social worker use observed between 2017 and 2019. Consistent with previous research, family doctors were found to be the most commonly accessed mental health care provider, reaffirming the central role of primary care in Canada’s mental health system.^5,18^ Given the high rates and numbers of people who seek mental health care from family doctors it is important that family doctors are trained in assessment and treatment of mental health.^19^ Access to psychologists and social workers increased in recent years, particularly between 2017 and 2019, which may reflect changing public attitudes toward mental health, reduced stigma, and increased availability of services.^20–22^

Logistic regression analyses from the 2019–2020 CCHS cycle indicate significant differences in access to providers across different demographic groups. Women had higher odds of seeking support from family doctors and social workers compared to men, aligning with prior literature showing greater mental health service use among women.^23^ Age emerged as a significant predictor of provider type with older adults more likely to seek care from family doctors and younger individuals more likely to access psychologists and social workers. This trend may reflect generational differences in mental health literacy, familiarity with provider types, and access pathways.^24,25^ Additionally, mental health services delivered by psychologists and social workers are not universally covered by provincial health plans in Canada, and younger individuals may access these providers through post-secondary institutions, school-based programs, or employee insurance plans. The rise in mental health awareness among younger generations, combined with greater comfort in discussing psychological issues, may also increase their likelihood of seeking specialized care.^26,27^ In contrast, older adults may be more affected by stigma surrounding mental illness and thus prefer family doctors who can address mental and physical health simultaneously in a less stigmatizing setting.^25^

Income and education were associated with distinct patterns of provider use. Individuals with higher education were more likely to access psychologists, potentially reflecting the private- pay nature of these services and barriers related to insurance coverage. In contrast, individuals with lower income and education levels were more likely to see family doctors or social workers, who are often covered under public or community-based services. These patterns highlight ongoing structural inequities in access to mental health care, where the ability to pay often determines the type and intensity of support received.^28,29^ These findings support the need for publicly funded access to a broader range of providers to ensure that individuals are matched to appropriate services regardless of ability to pay.

Self-perceived mental and general health status were also associated to provider use. Individuals who rated their mental health as poor had significantly higher odds of accessing all four types of mental health providers. However, the findings also reveal potential disparities in access as those who were not visible minorities had higher odds of accessing family doctors and non-Indigenous individuals were more likely to access psychologists. These results align with existing literature documenting racial and cultural inequities in mental health service access in Canada, driven by factors such as discrimination, mistrust of providers, culturally unsafe care, and underrepresentation of racialized and Indigenous practitioners.^30–33^ These findings underscore the importance of promoting equitable mental health services and support community-based models of care that are grounded in trust and cultural responsiveness.

These findings reveal an inequitable mental health care system in Canada, where sociodemographic factors such as age, gender, income, education, and racial identity shape not only access but also the type of provider individuals are able to see. While need-based access patterns were evident there are clear structural and systemic barriers that limit equitable engagement with care. These disparities reinforce the importance of expanding publicly funded mental health services beyond primary care and ensuring broader access to psychologists and social workers. As Canada continues to build toward a more integrated and accessible mental health system, efforts must prioritize reducing financial, geographic, and cultural barriers to ensure that all individuals can access the mental health care they need. Future research should explore the intersection of provider preferences, treatment experiences, and long-term outcomes to further inform changes to the mental health care system.

## Limitations

This study has several limitations. First, mental health service use was based on self-report and may be subject to recall or reporting biases. Second, only service use within the past year was captured without accounting for frequency, duration, or perceived adequacy of care. Third, the regression models only used cross-sectional data from 2019–2020. This is due to an inability to combine data from different cycles together as predictor variables were measured differently across various cycles of the survey. This means that logistic regression results are not able to describe longer-term patterns. Additionally, service use was measured separately for each provider type, without information on concurrent or integrated care.

## Conclusion

This study provides national evidence on who accesses which mental health providers in Canada and how that varies over time. While primary care remains a central entry point for mental health services, the growing role of psychologists and social workers may reflect changes in public demand and service availability. However, significant disparities persist particularly in access to privately funded services. These findings underscore the need for policy efforts to improve equity in mental health care delivery across provider types and population groups.

## Data Availability

The data that support the findings of this study are provided by Statistics Canada microdata files.

## Data Access

The data that support the findings of this study are provided by Statistics Canada microdata files.

## Acknowledgements

N/A

